# Bayesian estimation of IVW and MR-Egger models for two-sample Mendelian randomization studies

**DOI:** 10.1101/19005868

**Authors:** Okezie Uche-Ikonne, Frank Dondelinger, Tom Palmer

## Abstract

We present our package, `mrbayes,` for the open source software environment R. The package implements Bayesian estimation for IVW and MR-Egger models, including the radial MR-Egger model, for summary-level data in Mendelian randomization analyses. Users have the option of fitting the models using either JAGS or Stan software packages. We have implemented a choice of prior distributions for the model parameters, namely; weakly informative, non-informative, a joint prior for the MR-Egger model slope and intercept, and an informative prior (pseudo-horseshoe prior), or the user can specify their own prior. Similar prior distributions are included using the Stan software with the exception of a user-defined prior. We include We show how to use the package through an applied example investigating the causal effect of BMI on acute ischemic stroke. In future work, we plan to provide functions for Multivariable MR estimation.

## Introduction

Observational epidemiology is limited by possible bias due to unmeasured confounders, reverse causation and other problems. Mendelian randomization (MR) is a method of testing and estimating causal effects for the aetiology of diseases^1^. MR uses genetic variants as instrumental variables related to a modifiable phenotype to estimate a causal effect of the phenotype on a disease outcome. By including multiple instruments, we can increase power for hypothesis testing. The trade-off with this approach is the risk of violating the second and third instrumental variable assumptions due to horizontal pleiotropy.

Genome wide association studies (GWAS) provide many potential instruments, and we can obtain summary-level datasets for MR analyses. For two-sample MR, we get the values of instrument-phenotype associations and instrument-outcome associations from different samples.

The inverse variance weighted (IVW) model estimates the causal effect for multiple independent instruments in summary data. However, in the presence of pleiotropy its estimates are biased. Methods have been derived which estimate causal effects that are robust to pleiotropy; such as the MR-Egger model.^2^ The MR-Egger model relies on its InSIDE (Instrument strength is independent of direct effect) assumption. The MR-Egger model has recently been adapted with its radial formulation which has the advantage of viewing the instrument specific IV estimates in a radial plot and the IVW model is its sub-model^3^.

Extending MR analysis with Bayesian estimation allows specification of prior distributions on the model parameters. Several authors have considered Bayesian estimation of MR models including assessing different model parameterisations and Bayesian model averaging.^4,5^. Bayesian methods have also been applied towards dependant instruments and invalid instruments^6^. The use of weakly informative prior distributions in the MR-Egger model has been shown to have good coverage properties^8^.

This paper introduces our `mrbayes` R package that implements Bayesian estimation of the IVW, MR-Egger, and Radial MR-Egger models. The models are estimated using Markov chain Monte Carlo(MCMC) methods through an R interface to the JAGS and Stan software (using the `rjags` and `rstan` packages).^9,10^ Our package includes some specified prior distributions; non-informative, weakly informative, a shrinkage prior on the causal effect estimate (Pseudo-Horseshoe prior), and a joint prior on the intercept and causal effect estimate in the MR-Egger and radial MR-Egger models. The package also allows users to specify their own prior distributions within JAGS software.

In the next section, we briefly introduce the models included in our package. We then introduce the features of our `mrbayes` package, and we show how to use the package in an applied example. The supplementary material includes additional points relating to the methodology and examples.

### Methods

Equations (1), (2) and (3) denote the IVW, MR-Egger and Radial MR-Egger models respectively. Please see the supplementary material for additional explanation of the models.

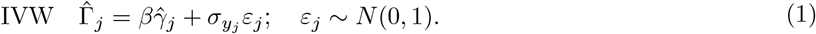

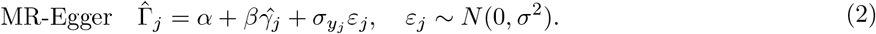

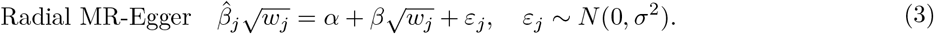

#### Prior Distributions

Table 1 gives a summary of the default prior distributions for the parameters in the mrbayes package. The supplementary material gives additional details about the prior distributions for the model including a covariance between intercept and slope in the MR-Egger models.

**Table 1:** Formula for default prior models in mrbayes. For functions in IVW model, there is no *α* parameter

| Model | Priors |
| --- | --- |
| Uninformative Priors | $\alpha \sim N(0, 1000), \beta \sim N(0, 1000), \sigma \sim U(0.0001, 10)$ |
| Weakly-Informative Priors | $\alpha \sim N(0, 1), \beta \sim N(0, 1), \sigma \sim U(0.0001, 10)$ |
| Pseudo-Horseshoe Priors | $\alpha \sim N(0, 1), \beta \sim C(0, 1), \sigma \sim IG(0.5, 0.5)$ |
| Joint Priors | Please see supplementary material |

### Implementation

Our mrbayes package provides the following functions:

- `mr_format`, a function for setting up the summary-level dataset for analysis.

The functions that use JAGS/Stan software are;

- `mr_ivw_rjags/mr_ivw_stan`, a function for estimating causal effects using the Bayesian IVW model, with a choice of prior distributions;
- `mr_egger_rjags/mr_egger_stan`, a function for estimating causal effects through the Bayesian MR-Egger model, with a choice of prior distributions;
- `mr_radialegger_rjags/mr_radialegger_stan`, a function for performing Bayesian analysis under the radial formulation of MR-Egger.

The package allows users:

- to specify custom prior distributions for the estimate of the causal effect (`betaprior`) and optionally for the residual standard error (`sigmaprior`) for the MR-Egger models (original and radial). This option is only for `_rjags` functions, the prior distributions are written in the JAGS syntax. For more information on how to specify prior distributions see page 34 of JAGS manual;^11^
- to choose a random seed for reproducible results and to choose the number of chains for MCMC, each chain should have a different seed;
- to set parameter `rho`, the correlation coefficient between the average pleiotropic effect and causal estimate. This option is only relevant when using the *joint* prior method;
- to plot the posterior density and investigate the MCMC diagnostics.

The package also includes two summary-level datasets containing:

- 185 SNPs with multiple instrument-phenotype associations for low-density lipoprotein cholesterol (LDL-c), while the instrument-outcome associations for coronary heart disease (CHD);^12^
- 14 SNPs with instrument-phenotype associations of body mass index (BMI) and instrument-outcome associations of insulin resistance.^13^

The next section shows an applied example with instructions in r code chunks on how to use the package.

### Example: Investigating the effect of BMI on acute ischemic stroke

We demonstrate the package using the motivating example by zhao and colleagues 14 which is also the example dataset in `mr.raps` for estimating the causal effect of body mass index (BMI) on acute ischemic stroke (AIS). We estimate the causal effect on the summary-level dataset with GWAS p-value threshold as *p ≤* 5 × 10^−8^ and when all the instruments are included. We apply the prior distributions in table 1 and compare with the frequentist model. Firstly, we load the package into our R session (See the Availability section for installation instructions).

~~~
library(mrbayes)
~~~

The next stage involves the setting up the dataset by using the `mr_format()` function. In our illustration the datasets are denoted as `bmi1` and `bmi2`;

~~~
dat <-
mr_format(
rsid = bmi1$SNP,
xbeta = bmi1$beta.exposure,
ybeta = bmi1$beta.outcome,
xse = bmi1$se.exposure,
yse = bmi1$se.outcome
)
dat2 <-
mr_format(
rsid = bmi2$SNP,
xbeta = bmi2$beta.exposure,
ybeta = bmi2$beta.outcome,
xse = bmi2$se.exposure,
yse = bmi2$se.outcome
)
~~~

We show the R syntax to estimate the Bayesian models using the default prior distributions. The code chunk below describes the syntax for each prior distributions for JAGS and Stan software. For the joint prior we assume the correlation between the intercept and slope is 0.5.

~~~
*## weakly informative prior*
*### JAGS*
vague_ivw <-
mr_ivw_rjags(
dat,
prior = “weak”,
seed = c(123456, 456789, 342564),
n.chains = 3
)
vague_mregger <-
mr_egger_rjags(
dat,
prior = “weak”,
seed = c(123456, 456789, 342564),
n.chains = 3
)
vague_radmregger <-
mr_radialegger_rjags
(
dat,
prior = “weak”,
seed = c(123456, 456789, 342564),
n.chains = 3
)
*### Stan*
vague_ivw_stan <-
mr_ivw_stan(dat,
prior = 2,
seed = 12345,
n.chains = 3)
vague_mregger_stan <-
mr_egger_stan(dat,
prior = 2,
seed = 12345,
n.chains = 3)
vague_radmregger_stan <-
mr_radialegger_stan(dat,
prior = 2,
seed = 12345,
n.chains = 3)
*## Default shrinkage prior*
*### JAGS*
pseudo_ivw <-
mr_ivw_rjags(
dat,
prior = “pseudo”,
seed = c(123456, 456789, 342564),
n.chains = 3
)
pseudo_mregger <-
mr_egger_rjags(
dat,
prior = “pseudo”,
seed = c(123456, 456789, 342564),
n.chains = 3
)
pseudo_radmregger <-
mr_radialegger_rjags(
dat,
prior = “pseudo”,
seed = c(123456, 456789, 342564),
n.chains = 3
)
*### Stan*
pseudo_ivw_stan <-
mr_ivw_stan(dat,
prior = 3,
seed = 12345,
n.chains = 3)
pseudo_mregger_stan <-
mr_egger_stan(dat,
prior = 3,
seed = 12345,
n.chains = 3
pseudo_radmregger_stan <-
mr_radialegger_stan(dat,
prior = 3,
seed = 12345,
n.chains = 3)
*## Joint Prior*
*### JAGS*
joint_mregger <-
mr_egger_rjags(
dat,
prior = “joint”,
rho = 0.5,
seed = c(123456, 456789, 342564),
n.chains = 3
)
joint_radmregger <-
mr_radialegger_rjags(
dat,
prior = “joint”,
rho = 0.5,
seed = c(123456, 456789, 342564),
n.chains = 3
)
*### Stan*
joint_mregger_stan <-
mr_egger_stan(dat,
prior = 4,
seed = 12345,
n.chains = 3)
joint_radmregger_stan <-
mr_radialegger_stan(dat,
prior = 4,
seed = 12345,
n.chains = 3)
~~~

**Table 2:**
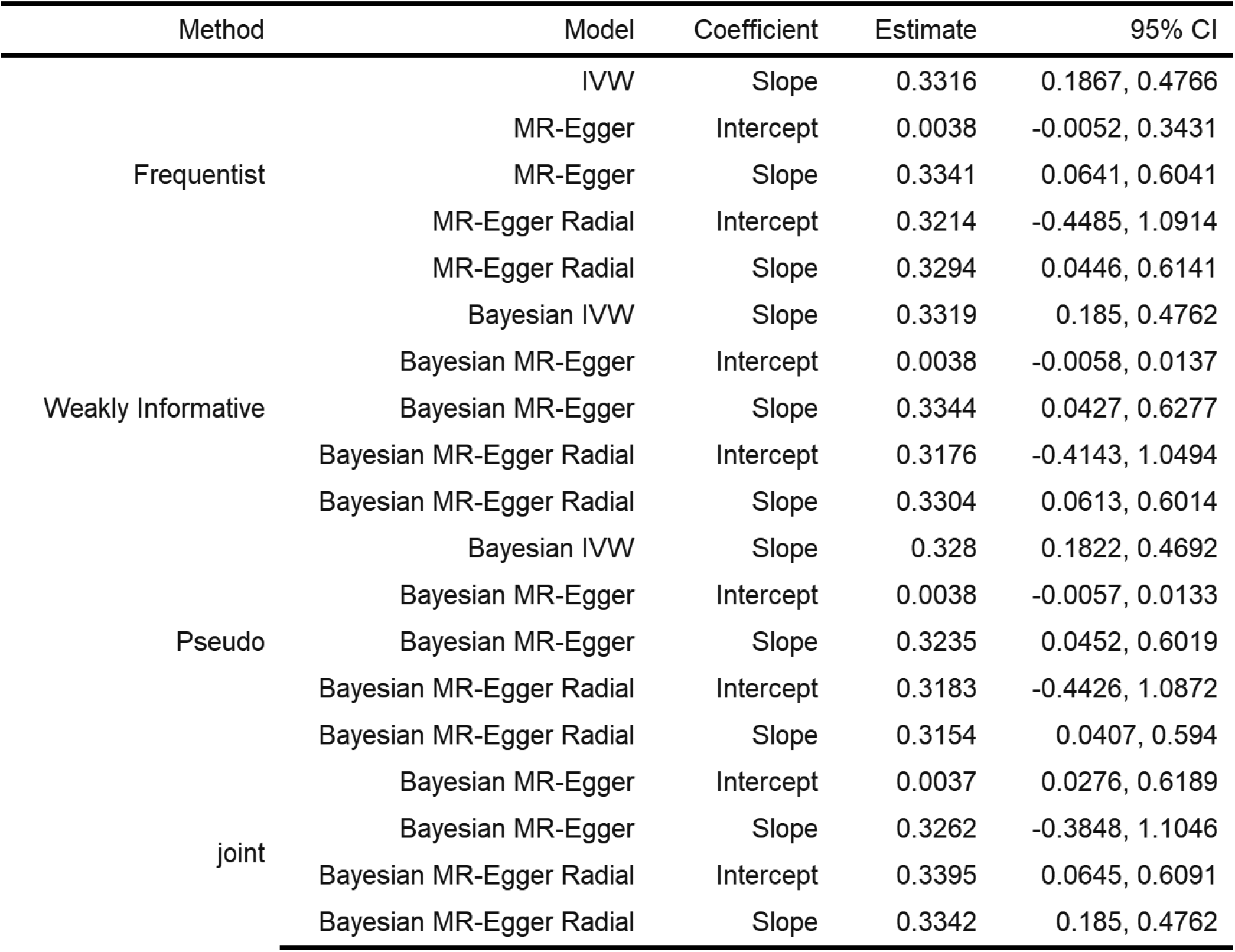
Estimates from summary-level data (GWAS p-value threshold). CI in the column indicates Confidence/Credible Interval The estimates and credible intervals from JAGS and Stan models are similar, we include estimates from JAGS in tables 2 and 3. The estimates derived from the models are seen in Tables 2 and 3 (dataset including all the instruments). In summary, estimates from the prior distributions for the MR-Egger model are consistent, figure 1 shows a graphical summary.

**Table 3:** Estimates from summary-level dataset when all instruments are included. CI in the column indicates Confidence/Credible Interval

| Method | Model | Coefficient | Estimate | 95% CI |
| --- | --- | --- | --- | --- |
| Frequentist | IVW | Slope | 0.3173 | 0.2117, 0.4229 |
|  | MR-Egger | Intercept | 0.0001 | -0.004, 0.3214 |
|  | MR-Egger | Slope | 0.3173 | 0.0998, 0.5348 |
|  | MR-Egger Radial | Intercept | 0.0015 | -0.32, 0.323 |
|  | MR-Egger Radial | Slope | 0.3173 | 0.0981, 0.5364 |
| Weakly Informative | Bayesian IVW | Slope | 0.3175 | 0.2104, 0.4226 |
|  | Bayesian MR-Egger | Intercept | 0.0001 | -0.004, 0.0042 |
|  | Bayesian MR-Egger | Slope | 0.319 | 0.1043, 0.5396 |
|  | Bayesian MR-Egger Radial | Intercept | 0.0007 | -0.3203, 0.3228 |
|  | Bayesian MR-Egger Radial | Slope | 0.319 | 0.1056, 0.5396 |
| Pseudo | Bayesian IVW | Slope | 0.316 | 0.2124, 0.4194 |
|  | Bayesian MR-Egger | Intercept | 0.0001 | -0.0039, 0.0042 |
|  | Bayesian MR-Egger | Slope | 0.3095 | 0.0897, 0.5284 |
|  | Bayesian MR-Egger Radial | Intercept | 0.0025 | -0.3105, 0.3208 |
|  | Bayesian MR-Egger Radial | Slope | 0.3094 | 0.0973, 0.5215 |
| joint | Bayesian MR-Egger | Intercept | 0.0001 | 0.1002, 0.5318 |
|  | Bayesian MR-Egger | Slope | 0.3184 | -0.329, 0.3092 |
|  | Bayesian MR-Egger Radial | Intercept | -0.0013 | 0.0956, 0.5404 |
|  | Bayesian MR-Egger Radial | Slope | 0.3222 | 0.2104, 0.4226 |

**Figure 1:**
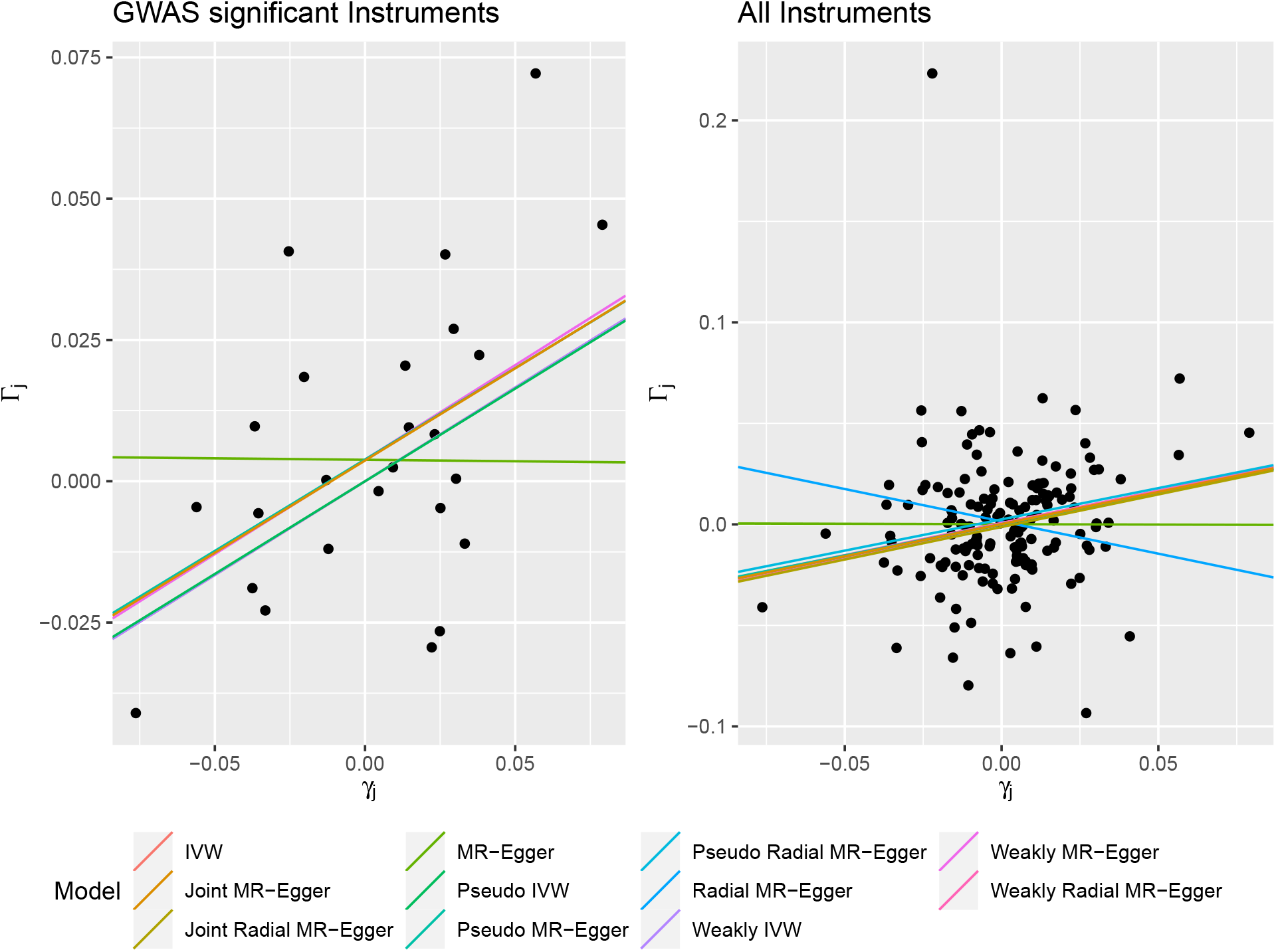
BMI and AIS

## Conclusion

We present an R package, mrbayes, to perform Bayesian estimation of the IVW and MR-Egger models implemented through the JAGS and Stan software. In our example, we demonstrated the use of several different prior distributions to estimate the causal and average pleiotropic effects from these models.

There are several R packages providing functions for MR analyses. The `MendelianRandomization` and `TwoSampleMR` packages implement various two-sample MR methods.^15,16^ The RadialMR R package implements the radial MR models and visualization of instruments through radial plots.^3,17^ Bayesian methods have not gained popularity in applied studies due to availability of a user-friendly software^18^. Our package complements previous MR packages by offering a Bayesian perspective with the choice of four prior distributions for the causal effect; non-informative, weakly informative, pseudo-horseshoe, and a joint prior distribution for the MR-Egger model’s intercept and slope.

In a Bayesian analysis the prior distributions can have an important impact upon the final parameter estimates. Hence in the mrbayes package we offer a choice of prior distributions. In future work, we plan to provide functions for multivariate models.

## Data Availability

Publicly available summary level data

## Availability

The package is freely available, under the MIT license, on GitHub here https://github.com/okezie94/mrbayes.

It can be installed in R using the following commands.

\# *install.packages(*“*remotes*”*) # uncomment if remotes not installed* remotes::install_github(okezie94/mrbayes”)

There is a website of the package helpfiles at https://okezie94.github.io/mrbayes/.

## References

1. Davey Smith G, Ebrahim S. ‘Mendelian randomization’: can genetic epidemiology contribute to understanding environmental determinants of disease? International Journal of Epidemiology. Oxford University Press; 2003;32(1):1–22.

2. Bowden J, Davey Smith G, Burgess S. Mendelian randomization with invalid instruments: effect estimation and bias detection through Egger regression. International Journal of Epidemiology [Internet]. 2015 Jun;44(2):512–525. Available from: https://dx.doi.org/10.1093/ije/dyv080

3. Bowden J, Spiller W, Del Greco M F, et al. Improving the visualization, interpretation and analysis of two-sample summary data Mendelian randomization via the Radial plot and Radial regression. International Journal of Epidemiology [Internet]. 2018 Jun;47(4):1264–1278. Available from: https://doi.org/10.1093/ije/dyy101

4. Jones E, Thompson J, Didelez V, Sheehan N. On the choice of parameterisation and priors for the Bayesian analyses of Mendelian randomisation studies. Statistics in Medicine. Wiley Online Library; 2012;31(14):1483–1501.

5. Burgess S, Thompson SG. Improving bias and coverage in instrumental variable analysis with weak instruments for continuous and binary outcomes. Statistics in Medicine. Wiley Online Library; 2012;31(15):1582–1600.

6. Shapland CY, Thompson JR, Sheehan NA. A bayesian approach to mendelian randomisation with dependent instruments. Statistics in medicine. Wiley Online Library; 2019;38(6):985–1001.

7. Berzuini C, Guo H, Burgess S, Bernardinelli L. A Bayesian approach to Mendelian randomization with multiple pleiotropic variants. Biostatistics [Internet]. 2018 Aug; Available from: https://dx.doi.org/10.1093/biostatistics/kxy027

8. Schmidt A, Dudbridge F. Mendelian randomization with Egger pleiotropy correction and weakly informative Bayesian priors. International Journal of Epidemiology. Oxford University Press; 2017;47(4):1217–1228.

9. Plummer M. Rjags: Bayesian graphical models using mcmc [Internet]. 2018. Available from: https://CRAN.R-project.org/package=rjags

10. Stan Development Team. RStan: the R interface to Stan [Internet]. 2018. Available from: http://mc-stan.org/

11. Plummer M. JAGS Version 3.3.0 user manual. Lyon, France: International Agency for Research on Cancer; 2012.

12. Do R, Willer CJ, Schmidt EM, et al. Common variants associated with plasma triglycerides and risk for coronary artery disease. Nature Genetics. Nature Publishing Group; 2013;45(11):1345.

13. Richmond R, Wade K, Corbin L, et al. Investigating the role of insulin in increased adiposity: Bi-directional Mendelian randomization study. *bioRxiv* [Internet]. Cold Spring Harbor Laboratory; 2017;155739. Available from: https://www.biorxiv.org/content/early/2017/06/28/155739

14. Zhao Q, Wang J, Hemani G, Bowden J, Small DS. Statistical inference in two-sample summary-data mendelian randomization using robust adjusted profile score. arXiv preprint arXiv:180109652. 2018;

15. Yavorska OO, Burgess S. MendelianRandomization: an R package for performing Mendelian randomization analyses using summarized data. International Journal of Epidemiology. Oxford University Press; 2017;46(6):1734–1739.

16. Hemani G, Zheng J, Elsworth B, et al. The MR-Base platform supports systematic causal inference across the human phenome. Elife. eLife Sciences Publications Limited; 2018;7:e34408.

17. Spiller W, Bowden J. Radial MR: A package for implementing radial inverse variance weighted and MR-Egger methods. [Internet]. 2019. Available from: https://github.com/WSpiller/RadialMR

18. Sheehan NA, Didelez V. Epidemiology, genetic epidemiology and mendelian randomisation: More need than ever to attend to detail. Human genetics. Springer; 2020;139(1):121–136.

